# Impacts of COVID-19 on glycemia and risk of diabetic ketoacidosis

**DOI:** 10.1101/2022.03.08.22272041

**Authors:** Anukriti Sharma, Anita D. Misra-Hebert, Arshiya Mariam, Alex Milinovich, Michael W. Kattan, Kevin M. Pantalone, Daniel M. Rotroff

## Abstract

**Background:** Reports indicate that COVID-19 may impact pancreatic function and increase type 2 diabetes (T2D) risk, although real-world COVID-19 impacts on HbA1c and T2D are unknown. We tested whether COVID-19 increased HbA1c, risk of T2D, or diabetic ketoacidosis (DKA).

**Methods:** We compared pre- and post-COVID-19 HbA1c, and risk of developing T2D in a large real-world clinical cohort of 8,755 COVID-19(+) patients and a matched control cohort of 11,998 COVID-19(-) patients. We investigated if DKA risk was modified in COVID-19(+) patients with type 1 diabetes (T1D) (N=704) or T2D (N=22,904), or by race and sex.

**Findings:** We observed a statistically significant, albeit clinically insignificant, HbA1c increase post-COVID-19 (all patients ΔHbA1c=0.06%, *P*<.001; with T2D ΔHbA1c=0.1%; *P*<.001), and no increase among COVID-19(-) patients (*P*>.05). COVID-19(+) patients were 40% more likely to be diagnosed with T2D compared to COVID-19(-) patients (*P*<.001) and 28% more likely to be diagnosed with T2D for the same HbA1c change as COVID-19(-) patients (*P*<.001). COVID-19(+) patients with T2D on insulin were 34% more likely to develop DKA compared to COVID-19(-) patients on insulin (*P*<.05), and COVID-19(+) Black patients with T2D displayed disproportionately increased DKA risk (HR:1.63, *P*=.007). There was no significant difference in DKA risk between COVID-19(+) and COVID-19(-) patients with T1D.

**Interpretation:** DKA risk is increased in T2D patients on insulin and in Black patients with T2D after COVID-19 infection.T2D risk is greater in COVID-19(+) patients for the same HbA1c increase in COVID-19(-) patients, indicating that T2D risk attributed to COVID-19 may be due to increased recognition during COVID-19 management.

**Funding:** No funding to report.

## Introduction

SARS-CoV-2 was recently demonstrated in two separate studies to infect pancreatic β cells *in vitro*^1,2^, raising concerns as to whether SARS-CoV-2 infection (COVID-19) may impact glycemic control^3^. Type 2 diabetes (T2D) is an established risk factor for COVID-19 severity and pre-infection hemoglobin A1c (HbA1c) was shown to be an important predictor of COVID-19 severity in multiple studies ^4-6^. Although HbA1c has been reported as an important risk factor for COVID-19, the impact of COVID-19 on HbA1c is unknown^6-8^. Previous studies have indicated that COVID-19 may increase HbA1c post-infection, but the cohorts were small and lacked COVID-19(-) controls for comparison ^9,10^. Furthermore, case studies of patients developing insulin-dependent diabetes and diabetic ketoacidosis (DKA) after COVID-19 have been reported^11,12^. Misra et al. demonstrated an increase in hospitalization for DKA in patients with T2D and a reduction in DKA hospitalization in patients with Type 1 diabetes (T1D), during the course of the COVID-19 pandemic ^13^. However, this study also lacked a negative control population to determine whether the risk was increased in patients that were infected with SARS-CoV-2 or was due to other factors. Here, we present the first evidence of COVID-19 impacts on HbA1c in a large, real-world clinical cohort of >8,500 patients by testing the following hypotheses 1) COVID-19 infection increases HbA1c, 2) COVID-19 infection increases risk of developing T2D, and 3) COVID-19 infection increases risk of DKA.

## Methods

### Study design and participants

We used the IRB approved COVID-19 data-registry established by Cleveland Clinic that contains data from 81,093 patients tested at the Cleveland Clinic who were COVID-19(+) between March 2020 and May 2021. In addition, the Cleveland Clinic registry included 153,034 matched patients that tested COVID-19(-) during the same time period. A 2:1 matched case-control approach was implemented to define a matched cohort of COVID-19(-) patients. Matching was performed based on location of the Cleveland Clinic facility, sex, race, age within five years, and in time slices of 14 days, but expanded to 21 days during the peak period of 11/8/2020-1/30/2021. Eligible controls were defined as patients that tested COVID-19(-) and had no record of previously testing COVID-19(+). This is analogous to control selection for a grouped-time, nested case-control study, although the status of patients who do not report for testing during any given time slice is unknown.

To evaluate COVID-19 impacts on HbA1c, we retrospectively analyzed the electronic health record and selected patients with an HbA1c recorded ≤12 months prior to their COVID-19 test, and an HbA1c recorded ≤12 months after their COVID-19 test. The final cohort meeting our inclusion criteria included 8,755 COVID-19(+) patients (3/2020-5/2021) and 11,998 COVID-19(-) patients (3/2020-4/2021). HbA1c from both pre- and post-COVID-19 test determined change in HbA1c in COVID-19(+) and COVID-19(-) groups. To investigate the impact of using up to 12 months between HbA1c testing and COVID-19 testing, we also analyzed two sub-cohorts for the same outcomes with a maximum time of 6 months and 3 months between HbA1c record date and the date of COVID-19 test. Using the 6 months criterion, 3,395 COVID-19(+) patients and 4,985 COVID-19(-) patients were included. Using 3 months criterion, 1,357 COVID-19(+) patients and 1,730 COVID-19(-) patients were included.

In order to evaluate the risk of DKA onset post-COVID-19 infection, we identified two sub-cohorts of patients with pre-existing T1D. Patients with a prior history of DKA at any time point prior to their COVID-19 test were excluded from this cohort. Patients were determined to have T2D based on a modified form of the eMERGE algorithm, as described in Kho et al.^14^ The eMERGE algorithm is designed to determine, with a high degree of specificity, which patients in an electronic health record have T2D based on a combination of ICD codes, medications, and laboratory values (e.g., blood glucose), and excludes patients with T1D. However, the original eMERGE algorithm utilized only ICD-9 codes, so the algorithm used here was modified to also include ICD-10 codes. T2D control patients were those that did not meet the eMERGE T2D criteria and had no history T1D. Patients were considered to have T1D as per the Phenotype Algorithm for T1D in the eMERGE Phase-IV program^15^. The T2D(+) cohort was further stratified into groups based on baseline insulin usage (yes or no), race and sex for DKA outcome assessment. Additionally, outcome of hyperosmolar hyperglycemic syndrome (HHS) was also assessed in T2D(+) cohort excluding patients with a prior history of HHS at any time prior to their COVID-19 test.

### Outcomes

The following outcomes were investigated for association with HbA1c in COVID-19(+) patients: T2D onset (based on eMERGE algorithm described above), hospitalization, admission to intensive care units (ICU), assisted breathing requirements, ventilation, and mortality. Additionally, DKA diagnosis was determined based on the ICD9/10 codes (ICD9:250.1x, ICD10:E10.1x, E11.1x). HHS was determined based on the ICD9/10 codes (ICD9:250.2x, ICD10:E11.0x).

### Statistical analyses

Statistical analyses were performed using R, v4.1.1^16^. Pre- and post-COVID-19 test HbA1c values were compared between patients 1) with and without T2D, and 2) with and without COVID-19(+) test results using paired Student’s t-tests. Logistic regression was employed to test for differences in new T2D diagnoses among COVID-19(+) and COVID-19(-) patients. HbA1c was tested for association with time-to-hospitalization, ventilation, assisted breathing, ICU admission, mortality, T2D onset using Cox proportional hazards (CoxPH) models. Mortality was used as a competing risk in all CoxPH models, except for when mortality was the outcome. Restricted cubic spline CoxPH models were used to investigate non-linear relationships between HbA1C and outcomes of interest^17^ (Supplemental Figures 3-6). Age, sex, race, BMI, pre-COVID-19 HbA1c, Charlson comorbidity score^18^, medications (including anti-diabetes therapies prior to infection and during the study window), prior comorbidities, the time intervals between the COVID-19 test and the pre-test HbA1c and the post-test HbA1c were used as model covariates for all analyses. For DKA and HHS onset, when not stratified by insulin use, we adjusted for insulin at baseline, in addition to the other covariates described above. Furthermore, Kaplan-Meier plots were also used to visualize the associations between DKA onset and COVID-19 test status in R using packages *survminer* and *survival* ^19,20^. All reported *P* have been adjusted for multiple comparisons using a false discovery rate approach and adjusted *P* < .05 were considered to be statistically significant ^21^.

## Results

### Clinical characteristics

We identified 6,322 patients with T2D among the 81,093 who were COVID-19(+) that also had corresponding HbA1c measurements for analysis. The patients in the COVID-19(+) and the matched COVID-19(-) cohorts (N=11,998) were similar in age, income, BMI, race and pre-COVID-19 HbA1c (Table 1). Similar observations were observed in the DKA and HHS cohort (Supplemental Table 1).

**Table 1.** Study participants and their evaluation.

| Characteristics | COVID-19 + |  |  | COVID-19 - |  |  |
| --- | --- | --- | --- | --- | --- | --- |
|  | Total COVID-19+ (N=8,755) | With T2D (N=6,322) | Without T2D (N=2,433) | Total COVID-19 - (N=11,998) | With T2D (N=5,404) | Without T2D (N=6,594) |
| Mean age, years | 60.0 | 61.6 | 56 | 59.2 | 63.7 | 55.6 |
| Mean BMI | 33.8 | 34 | 33.2 | 33 | 33.6 | 32.4 |
| Race: |  |  |  |  |  |  |
| Asian n (%) | 127 (1.5) | 96 (1.5) | 31 (1.3) | 229 (1.9) | 93 (1.7) | 136 (2.1) |
| Black or African American n (%) | 2,313 (26.4) | 1,785 (28.2) | 528 (21.7) | 2,895 (24.1) | 1,471 (27.2) | 1,424 (21.6) |
| Multiracial n (%) | 399 (4.6) | 273 (4.3) | 126 (5.2) | 502 (4.2) | 209 (3.9) | 293 (4.4) |
| White n (%) | 5,632 (64.3) | 3,972 (62.8) | 1,660 (68.2) | 7,944 (66.2) | 3,449 (63.8) | 4,495 (68.2) |
| Male n (%) | 4,056 (46.3) | 2,977 (47.1) | 1,079 (44.4) | 5,582 (46.5) | 2,672 (49.4) | 2,910 (44.1) |
| Median baseline HbA1c (%) | 6.3 | 6.7 | 5.5 | 6.0 | 6.7 | 5.7 |
| Median post-infection HbA1c (%) | 6.4 | 6.9 | 5.6 | 6.0 | 6.8 | 5.7 |
| Median income by zip code | 54,822 | 54,545 | 57,064 | 55,984 | 54,544 | 58,588 |
| **Outcomes** |  |  |  |  |  |  |
| Hospitalization n (%) | 2,903 (33.2) | 2,404 (38.0) | 499 (20.5) | NA | NA | NA |
| Ventilation n (%) | 480 (5.5) | 410 (6.5) | 70 (2.9) | NA | NA | NA |
| Assisted breathing n (%) | 1,878 (21.5) | 1,567 (24.8) | 311 (12.8) | NA | NA | NA |
| ICU admission n (%) | 645 (7.4) | 549 (8.7) | 96 (3.9) | NA | NA | NA |
| Mortality n (%) | 236 (2.7) | 212 (3.4) | 24 (0.9) | 402 (3.4) | 280 (5.2) | 122 (1.9) |
| **Comorbidities** |  |  |  |  |  |  |
| Current smoker n(%) | 566 (6.5) | 432 (6.8) | 499 (20.5) | 1,762 (14.7) | 788 (14.6) | 974 (14.8) |
| COPD n (%) | 972 (11.1) | 831 (13.1) | 70 (2.9) | 1,838 (15.3) | 1,154 (21.4) | 684 (10.4) |
| Asthma n (%) | 1,585 (18.1) | 1,193 (18.9) | 311 (12.8) | 2,753 (22.9) | 1,320 (24.4) | 1,433 (21.7) |
| Hypertension n (%) | 5,878 (67.1) | 4,649 (73.5) | 96 (3.9) | 8,826 (73.6) | 4,730 (87.5) | 4,096 (62.1) |
| Coronary artery disease n (%) | 1,582 (18.1) | 1,376 (21.8) | 24 (0.9) | 2,792 (23.3) | 1,823 (33.7) | 969 (14.7) |
| Heart failure n (%) | 1,126 (12.9) | 1,023 (16.2) | 499 (20.5) | 1,982 (16.5) | 1,441 (26.7) | 541 (8.2) |
| Cancer n (%) | 1,392 (15.9) | 1,083 (17.1) | 70 (2.9) | 2,676 (22.3) | 1,443 (26.7) | 1,233 (18.7) |
| History of transplant n (%) | 148 (1.7) | 144 (2.3) | 311 (12.8) | 277 (2.3) | 198 (3.7) | 79 (1.2) |
| Multiple sclerosis n (%) | 49 (0.6) | 33 (0.5) | 96 (3.9) | 92 (0.8) | 51 (0.9) | 41 (0.6) |
| **Connective tissue disease n (%)** | 368 (4.2) | 290 (4.6) | 24 (0.9) | 471 (3.9) | 253 (4.7) | 218 (3.3) |
| **IBD n (%)** | 143 (1.6) | 111 (1.8) | 499 (20.5) | 270 (2.3) | 152 (2.8) | 118 (1.8) |
| **Other immunosuppressive disease n (%)** | 1,251 (14.3) | 1,103 (17.5) | 70 (2.9) | 2,106 (17.6) | 1,413 (26.2) | 693 (10.5) |
| **Pregnant n (%)** | 7 (0.1) | 5 (0.1) | 311 (12.8) | 19 (0.2) | 2 (0.1) | 17 (0.3) |
| **Epilepsy n (%)** | 321 (3.7) | 260 (4.1) | 96 (3.9) | 582 (4.9) | 302 (5.6) | 280 (4.3) |
| **Charlson Comorbidity index (median)** | 5 | 4 | 3 | 5 | 5 | 4 |
| **Anti-diabetes therapies at baseline (%) \*** |  |  |  |  |  |  |
| **Insulin** | 51.7 | 50.2 | NA | 49.7 | 53.6 | NA |
| **Metformin hydrochloride** | 21.4 | 21.5 | NA | 21.3 | 19 | NA |
| **Glimepiride (sulfonylureas)** | 4.9 | 5.1 | NA | 5.2 | 4.8 | NA |
| **Glipizide ((sulfonylureas)** | 4.7 | 4.8 | NA | 4.9 | 4.9 | NA |
| **Sitagliptin phosphate (DPP4-inhibitors)** | 3.2 | 3.2 | NA | 3.7 | 3.7 | NA |
| **Liraglutide (GLP1RA)** | 1.8 | 1.9 | NA | 1.6 | 1.6 | NA |

*\*These are not mutually exclusive for patients. DPP-4 stands for dipeptidyl peptidase-4. Therapies with at least 1% prevalence are reported in this table.*

### COVID-19 impacts on post-infection HbA1c

A statistically significant, but clinically insignificant, increase in HbA1c was observed after testing COVID-19(+) (ΔHbA1c=0.06%, *P*<.001) whereas COVID-19(-) patients did not demonstrate an increase in HbA1c (ΔHbA1c=0.02%, *P*=.05) (Supplemental Figure 1a). HbA1c change between pre-and post-COVID-19 testing demonstrated a 0.08% greater HbA1c increase in COVID-19(+) patients compared to COVID-19(-) patients(*P*<.001) (Supplemental Figure 1b). After stratifying COVID-19(+) patients by T2D status, we still observed a significant increase post-infection in patients with and without T2D (ΔHbA1c=0.1%, *P*<.001) and (ΔHbA1c=0.1, *P*<.001), respectively. These changes were not clinically meaningful. Importantly, no significant increase was observed for COVID-19(-) cases after stratifying by T2D status (*P*>.05) (Figure 1). Restricting the pre-COVID-19 test HbA1c time interval from 12 months to 6 and 3 months resulted in no statistically significant differences between pre- and post-HbA1c data both overall and stratified by T2D status (*P*>.05).

**Figure 1.**
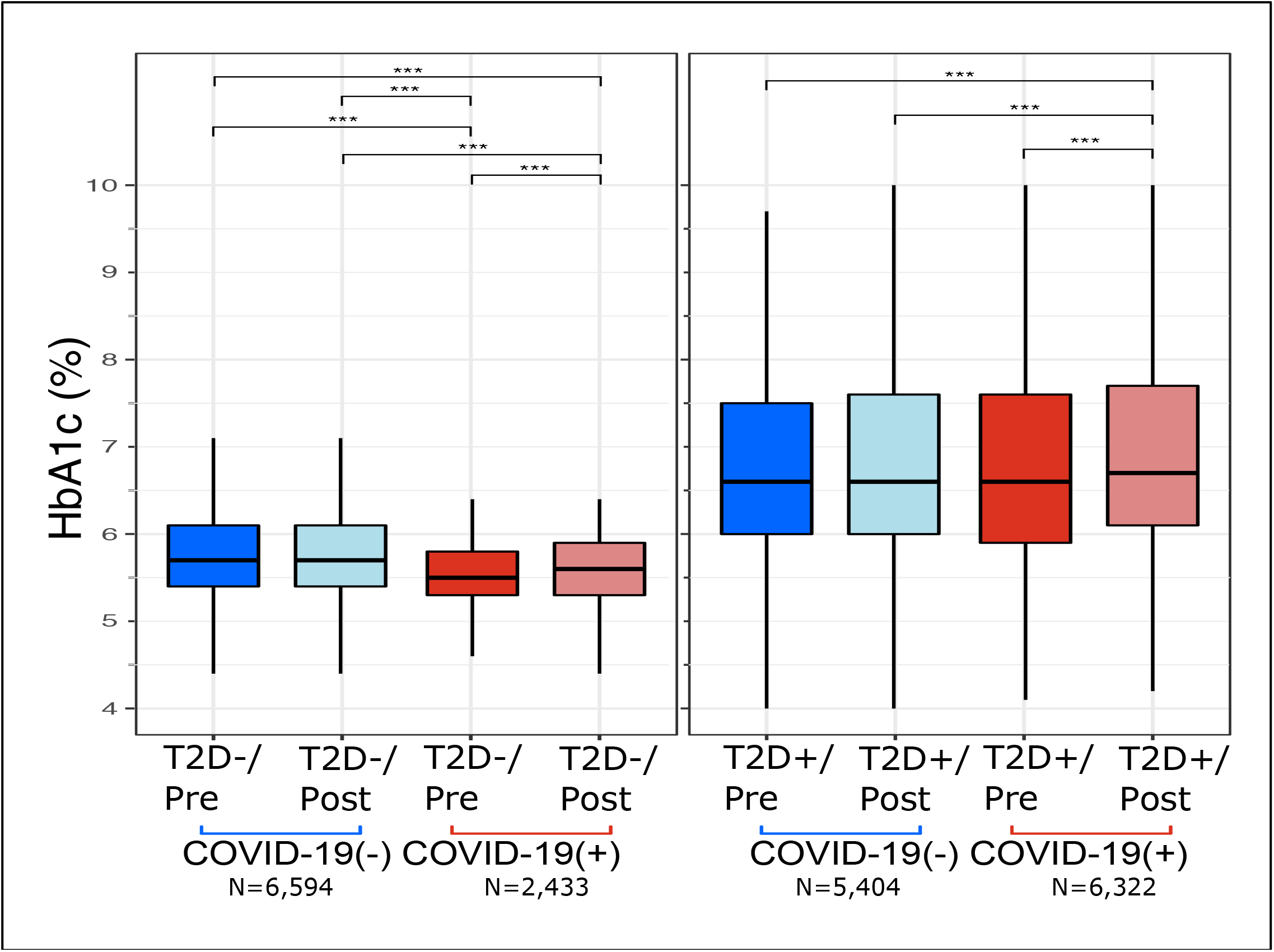
(A) HbA1c changes post-SARS-CoV-2 test in COVID-19(+) and COVID-19(-) patients, with and without type 2 diabetes (T2D). *, P<.05; **, P<.01; ***, P<.001

Compared to COVID-19(-) patients, a 0.09% increase in post-infection HbA1c was observed in COVID-19(+) patients with T2D (*P*<.001) (Supplemental Figure 1c). In patients without T2D, a clinically insignificant increase HbA1c of 0.05% in COVID-19(+) patients compared to COVID-19(-) patients (*P*<.001) was observed (Supplemental Figure 1c). Furthermore, the 0.09% HbA1c increase observed in patients with T2D was statistically greater than patients without T2D (0.05%) (*P*=.002). Comparable results were obtained after restricting the pre-COVID-19 test HbA1c time interval from 12 months to 6 and 3 months (data not shown).

### Pre-infection HbA1c and COVID-19 severity and mortality

Although well reported in the literature, we also investigated whether pre-COVID-19 HbA1c was associated with COVID-19 severity in our cohort (*i*.*e*., hospitalization, ventilation, assisted breathing, ICU admission). In the COVID-19(+) cohort of patients with T2D, we identified a significant positive association between pre-COVID-19 HbA1c and time to hospitalization (HR=1.07, *P*<.001), assisted breathing (HR=1.06, *P*<.001), and admission to ICU (HR=1.07, *P*=.07) (Supplementary Figure 1). We observed similar associations using restricted cubic spline non-linear CoxPH models (Supplemental Figure 2). Restricting the pre-COVID-19 test HbA1c time interval to 6-months and 3-months still resulted in significant associations with both hospitalization (HR_6mo_=1.08, *P*_6mo_<.001 & HR_3mo_=1.06, *P*_3mo_=.02) and assisted breathing (HR_6mo_=1.08, *P*_6mo_<.001 & HR_3mo_=1.07, *P*_3mo_=.02).

### COVID-19 infection and risk of type 2 diabetes

Pre-COVID-19 HbA1c was a significant risk factor for receiving a T2D diagnosis in COVID-19(+) patients (HR=4.08, *P*<.001) and COVID-19(-) patients (HR=1.34, *P*<.001), indicating that higher pre-COVID-19 test HbA1c increased risk of receiving a T2D diagnosis regardless of COVID-19 status (Figure 2A). Similarly, risk of T2D onset was higher in individuals with greater HbA1c increase in COVID-19(+) (HR=1.74, *P*<.001) and COVID-19(-) patients (HR=1.46, *P*<.001) (Figure 2D). However, COVID-19(+) patients were also more likely to receive a T2D diagnosis post-infection (N=326/2,433) compared to COVID-19(-) (N=380/6,594) patients (OR=1.4, *P*<.001). Interestingly, the likelihood of being diagnosed with T2D, per unit change in HbA1c, was also higher for COVID-19(+) patients than COVID-19(-) patients. Using the restricted cubic spline non-linear CoxPH models demonstrated similar association patterns (Supplemental Figure 3). Furthermore, restricting the pre-COVID-19 test HbA1c time interval from 12 months to 6 and 3 months resulted in similar significant trends for T2D onset for both COVID-19(+) as well as COVID-19(-) control group (data not shown).

**Figure 2.**
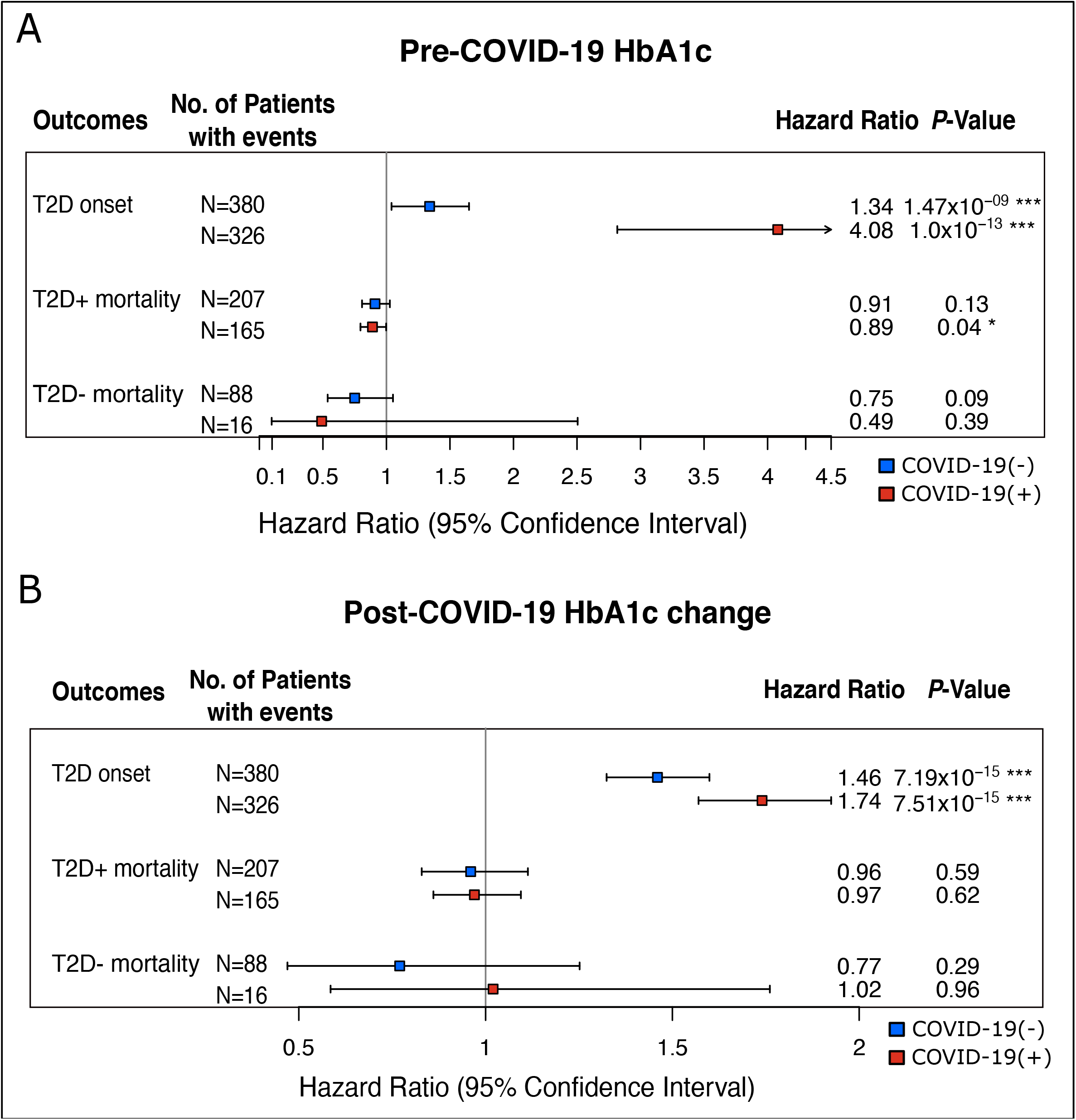
(A) Risk of T2D onset and mortality with baseline HbA1c and (B) change in HbA1c in COVID-19(+) and COVID-19(-) patients. Paired t-test comparisons were made for all.

### COVID-19 infection and risk of DKA and HHS

We evaluated the risk of DKA onset post-COVID-19 infection, in two sub-cohorts of patients with pre-existing T1D (n=701) and (T2D) (n=21,830) out of the total COVID-19(+) (n=81,093) and COVID-19(-) population (n=153,034). In patients with T1D, there was no statistically significant difference in risk of developing DKA between those that were COVID-19(+) and those that were COVID-19(-) (HR=0.978, *P*=.48) (Figure 3). However, patients with T2D that were COVID-19(+) had a 35% greater risk of DKA compared to patients with T2D that were COVID-19(-) (HR=1.35, *P*=.002) (Figure 3). Patients with T2D that were on insulin were at increased risk of DKA compared to patients not on insulin regardless of COVID-19 testing status (Figure 4A,B). However, stratification of patients with T2D by insulin usage, also revealed that COVID-19(+) patients with T2D on insulin had a 34% greater risk of DKA than COVID-19(-) patients on insulin (HR=1.34, *P*=.02) (Figure 3). The risk of DKA between patients with T2D that were not on insulin was not significantly different between those that were COVID-19(+) and those that were COVID-19(-) (HR=1.30, *P*=.29) (Figure 3). No significant associations were observed between HHS (N=163) and COVID-19 infection status in patients with T2D (N=22,966) (Supplementary Figure 7).

**Figure 3.**
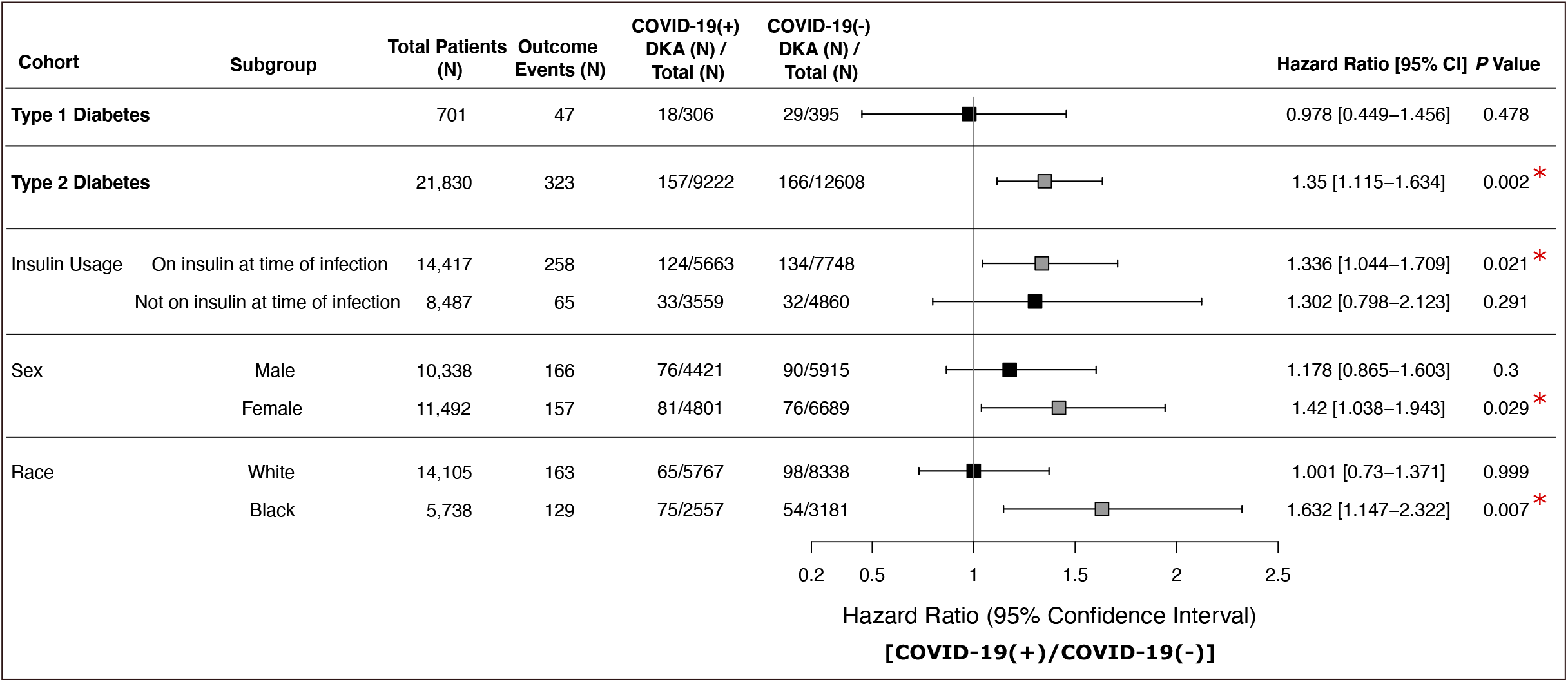
(A) Association of risk of Diabetic ketoacidosis (DKA) onset and COVID-19 test status in patients with pre-existing type 1 diabetes (+) and T2D(+) using coxPH models illustrated by forest plot. The T2D(+) cohort was further stratified based on baseline insulin usage, sex, and race. The patients with pre-existing DKA were removed from the T1D(-) and T2D(+) cohorts. The boxes in grey represent the significant associations (P<0.05). The column COVID-19(+) events represent the proportion of number of [COVID-19(+)/DKA(+)]/[COVID-19(+)/DKA(+/-)] events. Similarly, the column COVID-19(-) events represent the proportion of number of [COVID-19(-)/DKA(+)]/[COVID-19(-)/DKA(+/-)] events.

**Figure 4.**
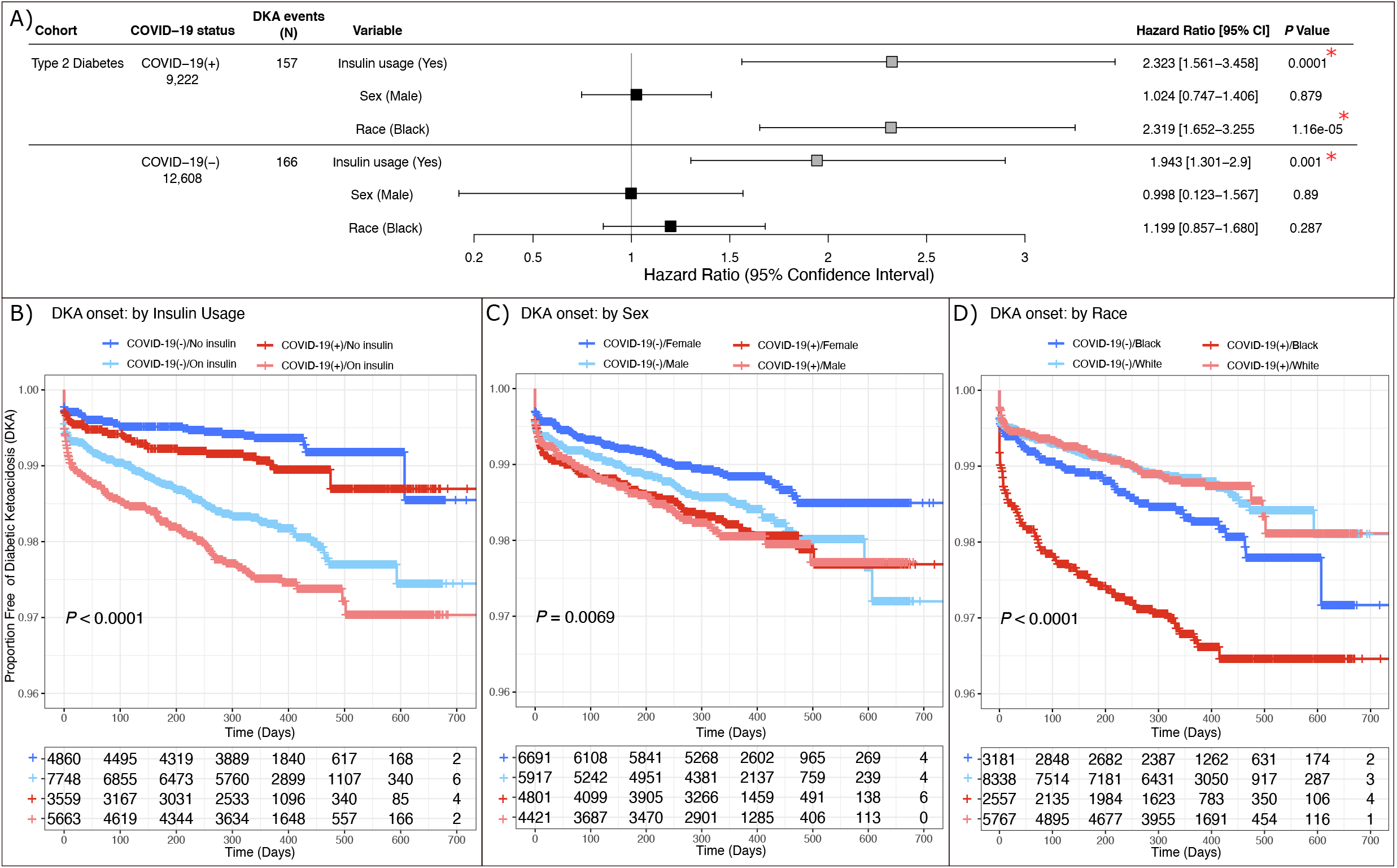
(A) Association of risk of Diabetic ketoacidosis (DKA) onset and insulin usage, sex and race in patients with pre-existing T2D who tested COVID-19(+). The boxes in grey represent the significant associations (P<0.05). Kaplan-Meier plots showing association between DKA onset and (B) insulin usage, (C) sex, and (D) race, in T2D(+) cohort of patients who tested COVID-19(+) as well as COVID-19(-).

When stratified by race, Black patients with T2D that were COVID-19(+) demonstrated a 63% greater risk of DKA compared to the Black patients with T2D that were COVID-19(-) (HR=1.63, *P*=.007) (Figure 3). There was no difference observed between the risk of DKA for White patients with T2D based on their COVID-19 testing result (HR=1.00, *P*=0.99) (Figure 3). In addition, the difference in DKA risk between Black and White patients with T2D that were COVID-19(-) was not significantly different (HR: 1.20, *P*=.29). However, in patients that were COVID-19(+), Black patients with T2D were more than twice as likely to be diagnosed with DKA compared to White patients with T2D (HR=2.32, *P*=1.16×10^−5^) (Fig. 4A).

There was no significant difference in DKA risk between males and females with T2D that were COVID-19(+) (HR=1.02, *P*=.88) or COVID-19(-) (HR=1.00, *P*=.89) (Figure 4A,C). The increase observed in males with T2D that were COVID-19(+) compared to those that were COVID-19(-) was not statistically significant (HR=1.80, *P*=.30). However, females with T2D demonstrated a significantly higher risk for DKA if COVID-19(+) (HR=1.42, *P*=.03) (Figure 3A). Therefore, although there was no difference in risk of DKA between males and females that were COVID-19(+) (Figure 4A), there was a relative increase in risk of females that were COVID-19(+) to those that were COVID-19(-) (Fig. 3,4A).

## Discussion

The impact of COVID-19 on HbA1c and glycemic impairment is still poorly understood. Yang et al. demonstrated that human pluripotent stem cell-derived and adult human pancreatic β cells were permissive to SARS-CoV-2 infection *in vitro* ^1^. This was further supported by findings that SARS-CoV-2 was capable of infecting human pancreatic islets *ex vivo* and impaired insulin secretion in an *in vitro* model system ^2^. Further raising concerns that COVID-19 might impair glycemic control, Montefusco et al. found that 46% of 551 patients hospitalized for COVID-19(+) were hyperglycemic, but this analysis lacked a COVID-19(-) population for comparison ^10^. A meta-analysis of 179 patients showed an increase in blood glucose in COVID-19(+) patients that failed to reach statistical significance^9^. Other forms of glycemic impairment have also been reported—a case study recently reported a patient that developed by autoantibody-negative insulin-dependent diabetes (type 1B subtype) that presented as DKA several weeks post-COVID-19 infection ^12^. A second case study of an otherwise healthy patient developing DKA while infected with SARS-CoV-2 was also reported, although the autoantibody status of this patient was not reported ^11^.

Although these studies suggest the potential for COVID-19 to disrupt glycemic response, no large controlled clinical studies have been conducted to determine the extent of this risk in patients diagnosed with COVID-19. This study improves upon limitations of previous studies by (1) utilizing a large cohort of 8,755 COVID-19(+) patients, (2) investigating pre- and post-infection HbA1c levels with respect to T2D status, and (3) including a large, matched cohort of 11,998 COVID-19(-) patients for comparison. Our findings indicate that there is an approximate 0.1% increase post-infection HbA1c among patients with and without T2D, which is statistically significant but clinically insignificant (Figure 1a). Although COVID-19(+) patients were 40% more likely to be diagnosed with T2D compared to COVID-19(-) patients (*P*<.001), we also found that COVID-19(+) patients were 274% more likely to get diagnosed with T2D for the same pre-COVID-19 test HbA1c and 28% more likely for the same increase in HbA1c after their COVID-19 test (Figure 1c) (*P*<.001). A possible explanation for this is that COVID-19(+) patients are receiving more intensive care that results in better identification of patients that have underlying T2D.

Interestingly, among patients with pre-existing T2D, we observed a 35% increased risk of developing DKA in COVID-19(+) patients compared to COVID-19(-) patients (*P*=.002). Importantly, this observation seems to be driven in part by patients that were on insulin at the time of their COVID-19 diagnosis (HR=1.34, *P*=.02), whereas the increase in DKA in patients with T2D that were not on insulin failed to reach significance (HR:1.30, *P*=.29) (Figure 4). Notably, COVID-19 infection did not increase DKA in patients with pre-existing T1D. A previous study found that hospital admissions for DKA increased compared to preceding years in patients with T2D during the COVID-19 pandemic but decreased for patients with T1D over the same time period ^13^. It has been hypothesized that the difference in DKA incidence between patients with T1D and T2D may be due to the training of patients with T1D to titrate insulin based on glucose levels, whereas patients with T2D may check their glucose levels more infrequently, resulting in higher insulin levels^13^. We also observed a higher risk of DKA onset in the Black COVID-19(+) patients with T2D compared to the Black COVID-19(-) patients with T2D (HR=1.632, *P*=.007), a relationship not observed in White patients (Figures 3,4). In addition, there was a relative increase in risk for DKA of females with T2D that were COVID-19(+) compared to females with T2D that were COVID-19(-) (HR: 1.42, *P*=.03), which was not observed in males (HR: 1.18, *P*=.30) (Figures 3, 4). These findings, in particular the findings of increased DKA in patients with T2D that are Black and those that are on insulin, which are consistent across studies, indicate that increased awareness is needed for DKA in these patient populations if infected with COVID-19.

As with any study, there are limitations that should be considered during the interpretation of the findings. To evaluate a large cohort of patients, we included patients with a HbA1c measurement for up to 12 months prior to their COVID-19 test. Although we adjusted for the time between HbA1c and COVID-19 testing, comorbid conditions, and medications during this period, it is possible that medication changes or other factors during the 12 months impacted HbA1c. To address this, we also evaluated the results using three- and six-month intervals, which yielded consistent results, indicating that these factors are unlikely to have a material impact on the analysis. Although consistent criteria were applied to the selection of COVID-19(+) and COVID-19(-) patients, HbA1c values were not available for many patients in the COVID-19 registry, and care was disrupted during the pandemic, which may bias the population available for inclusion in this study. Furthermore, this was a single-center study and patients may have received care at a different institution resulting in medical histories unavailable for this study. Additional studies at other sites and with longer follow-up data on HbA1c levels are needed to verify our results.

In conclusion, we observed a statistically significant, but clinically insignificant increase in HbA1c post-COVID-19 infection, and the increased likelihood of receiving a T2D diagnosis post-COVID-19 infection may be due to the increased care provided to these patients. Notably, in this large cohort of patients that were COVID-19(+), the risk of DKA was significantly higher in patients with pre-existing T2D overall and those on baseline insulin and those that are Black. Additional research is required to understand the pathogenesis of COVID-19 infection and DKA in patients with T2D, and additional clinical awareness is needed for DKA in these patient populations.

## Data Availability

Data related to this manuscript will be made available upon request in adherence with transparency conventions in medical research and through requests to the corresponding author.

## Author contributions

A.S. performed analyses and wrote the manuscript. Al.M. and Ar.M. extracted and curated data. K.M.P., A.M.H., and M.W.K. provided input into the study design, edited, and reviewed the manuscript. D.M.R. designed the study and wrote the manuscript.

## Acknowledgments

We would like to acknowledge the many people that worked to establish the Cleveland COVID-19 data-registry and their efforts to make this valuable resource available to investigators.

## Supplementary Material

**Supplementary Table 1:** Basic characteristics of patients in the cohorts of diabetic ketoacidosis DKA and Hyperosmolar hyperglycemic syndrome (HHS) onset with pre-existing type 2 diabetes (T2D).

| Characteristics | DKA/COVID-19(+) (N=157) | DKA/COVID-19(-) (N=166) | HHS/COVID-19(+) (N=75) | HHS/COVID-19(-) (N=88) |
| --- | --- | --- | --- | --- |
| Mean age, years | 59.5 | 60.5 | 62.6 | 61.8 |
| Mean BMI (SD) | 32.9 | 33.5 | 33.4 | 33.2 |
| Race (%): |  |  |  |  |
| Asian | 1.9 | 0.6 | 1.3 | 1.1 |
| Black or African American | 47.8 | 32.5 | 48 | 47.7 |
| Multiracial | 5.7 | 3.6 | 5.3 | 2.3 |
| White | 41.4 | 59.5 | 41.3 | 45.5 |
| Male (%) | 48.4 | 54.2 | 52 | 51.1 |
| Mean baseline HbA1c (SD) | 8.85 (2.2) | 8.46 (2.3) | 8.48 (2.6) | 8.47 (2.7) |

**Supplemental Figure 1.**
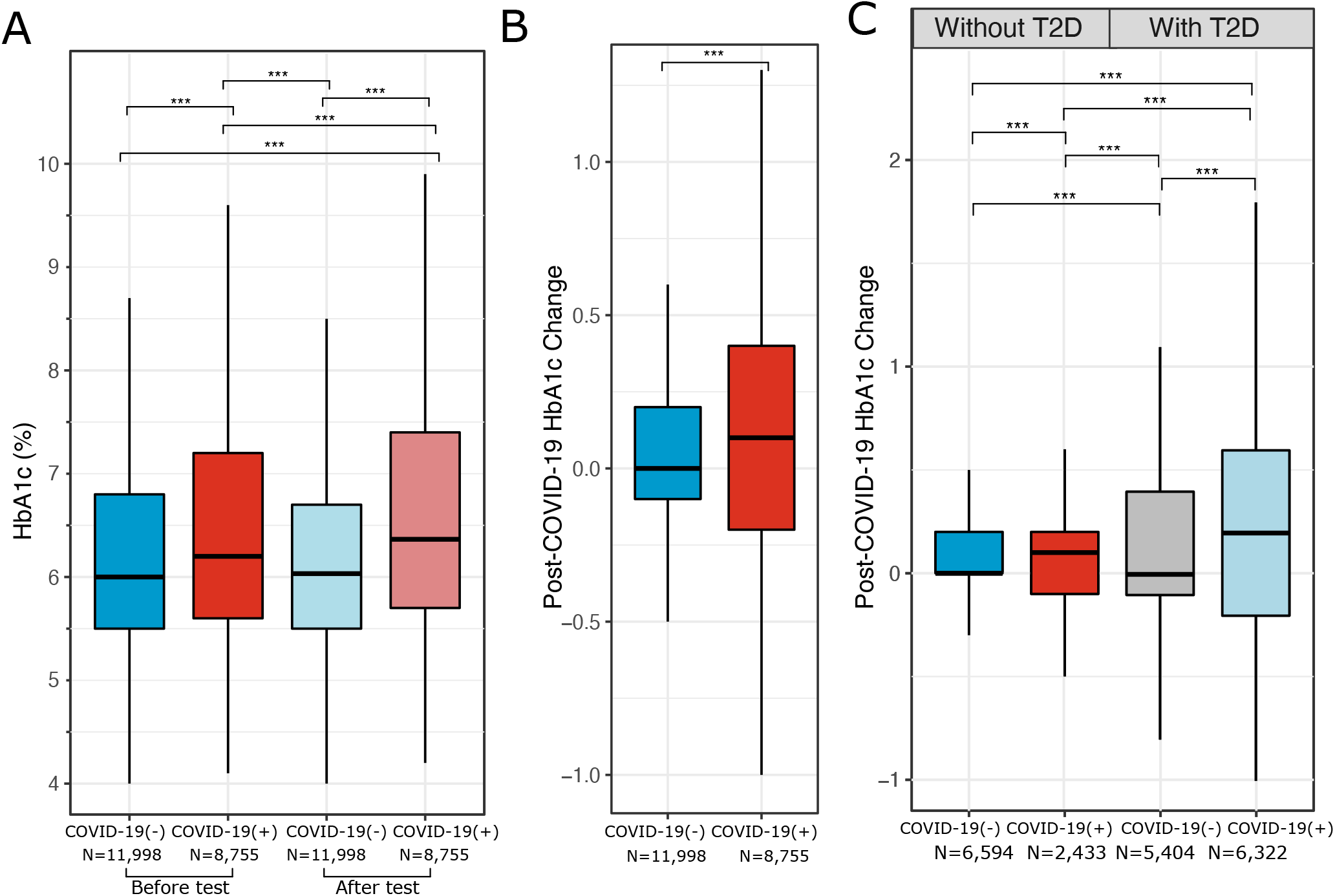
(A) HbA1c changes post-SARS-CoV-2 test in COVID-19(+) and COVID-19(-) patients overall without stratification by T2D status. Comparison of HbA1C changes between COVID-19(+) and COVID-19(-) cohorts (B) unstratified and (C) stratified by T2D status. Paired t-test comparisons were performed for all. **, P <0.01; ***, P < 0.001

**Supplementary Figure 2.**
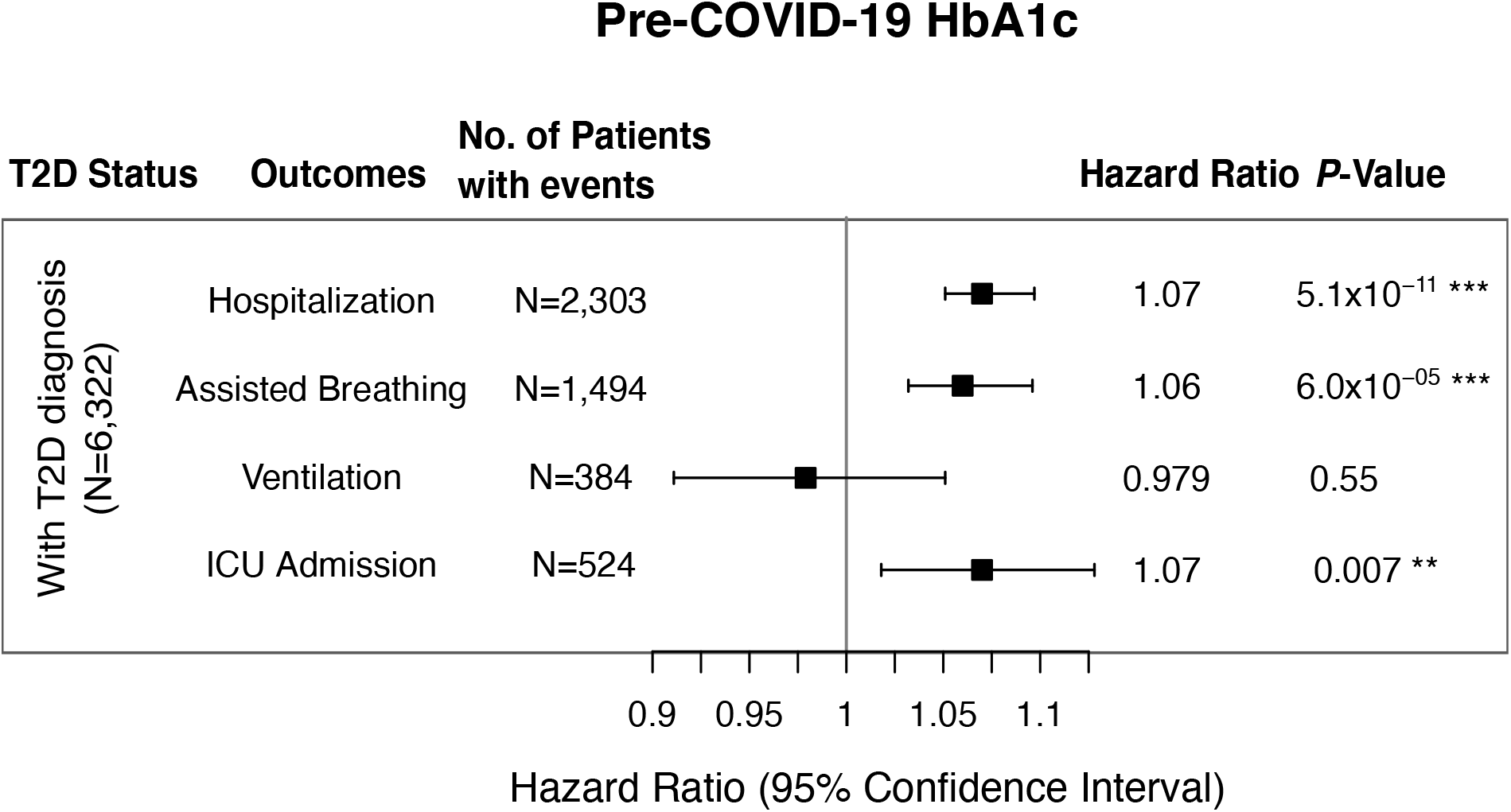
Risk of COVID-19 severity and mortality with baseline HbA1c in patients diagnosed with T2D.

**Supplementary Figure 3.**
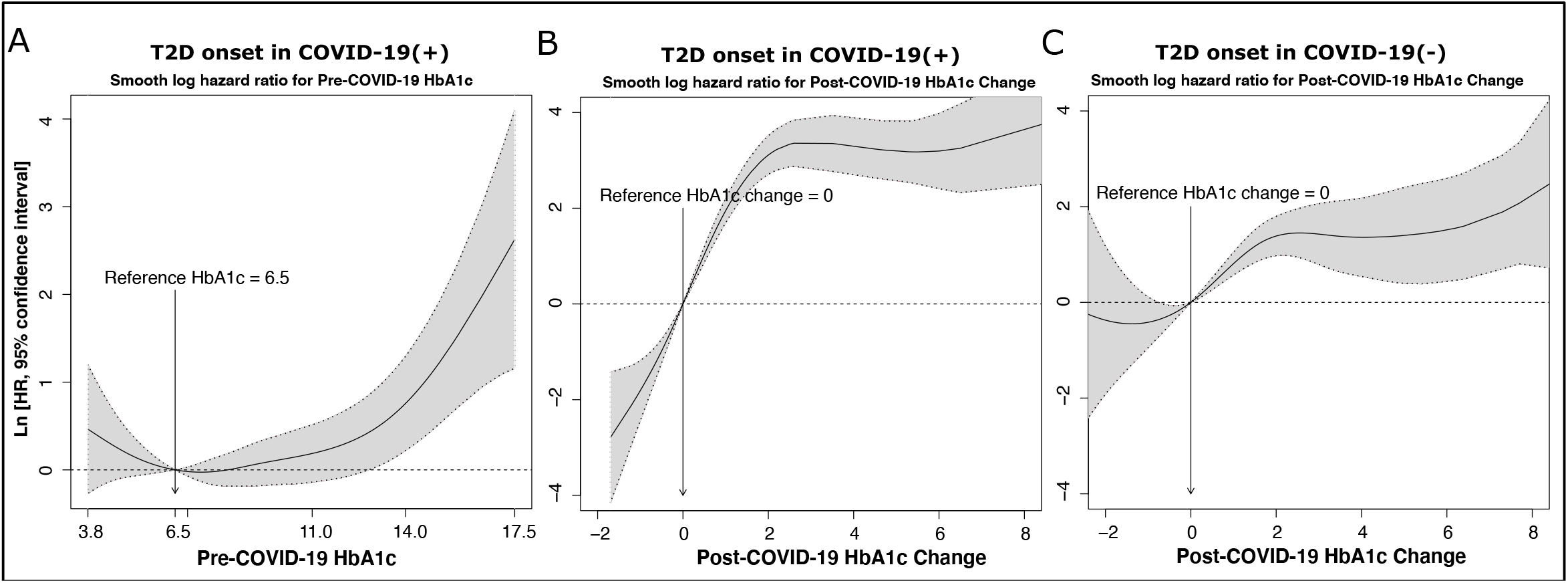
Non-parametric estimates of the dependence of risk of T2D diagnosis post-COVID-19 infection using restricted cubic splines in a Cox proportional hazards framework (A) on pre-COVID-19 HbA1c, (B) on post-COVID-19 HbA1c change in COVID-19(+) patients. (C) Dependence of T2D diagnosis post-COVID-19 testing on post-COVID-19 test HbA1c change in COVID-19(-) patients.

**Supplementary Figure 4:**
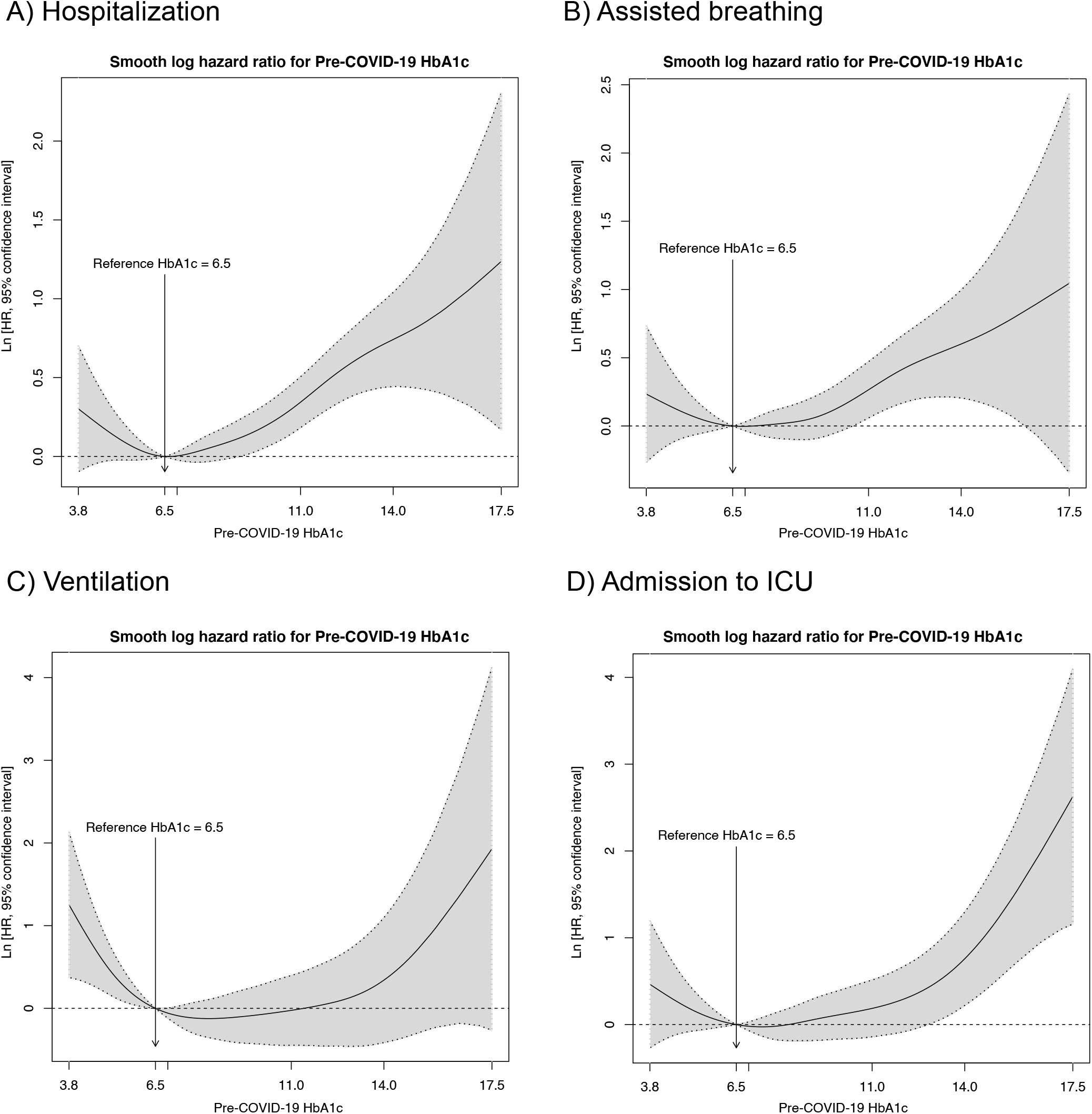
Non-parametric estimates of the dependence of all-time risk of (A) hospitalization, (B) assisted breathing, (C) ventilation, and (D) admission to ICU on baseline A1C among the COVID-19(+) patients with Type-2 Diabetes diagnosis. The plots were generated using restricted cubic splines implemented in Cox proportional hazard models using smoothHR package in R (2).

**Supplementary Figure 5:**
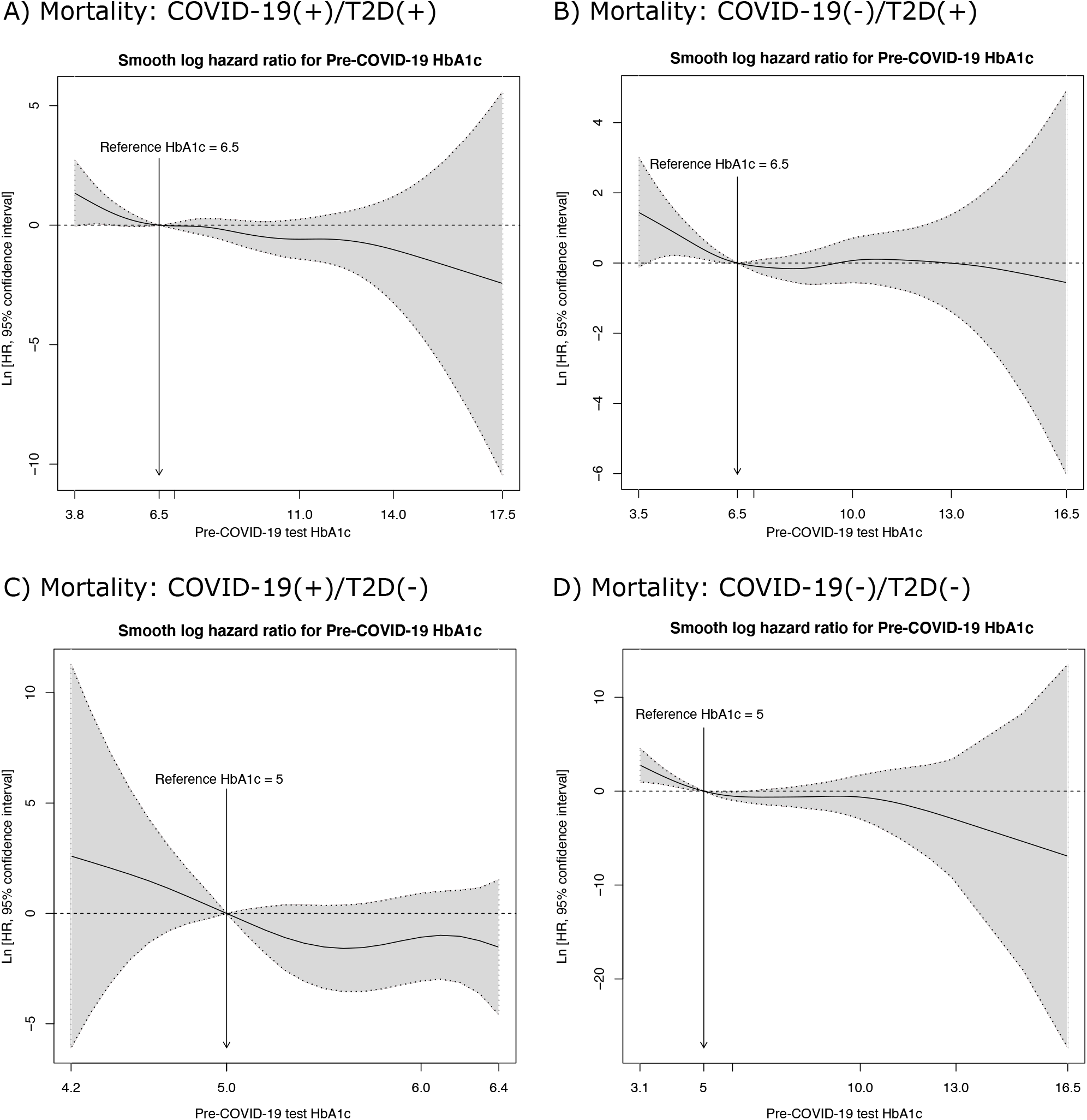
Non-parametric estimates of the dependence of risk of all-cause mortality on pre-COVID-19 test HbA1c in (A) COVID-19(+) patients with T2D diagnosis, (B) COVID-19(-) patients with T2D diagnosis, (C) COVID-19(+) patients without T2D diagnosis, and (D) COVID-19(-) patients without T2D diagnosis. The reference HbA1c values for T2D(+) cohort was set to 6.5 and for T2D(-) patients was set to 5. The plots were generated using restricted cubic splines (rcs) implemented in Cox proportional hazard models using smoothHR package in R (2).

**Supplementary Figure 6:**
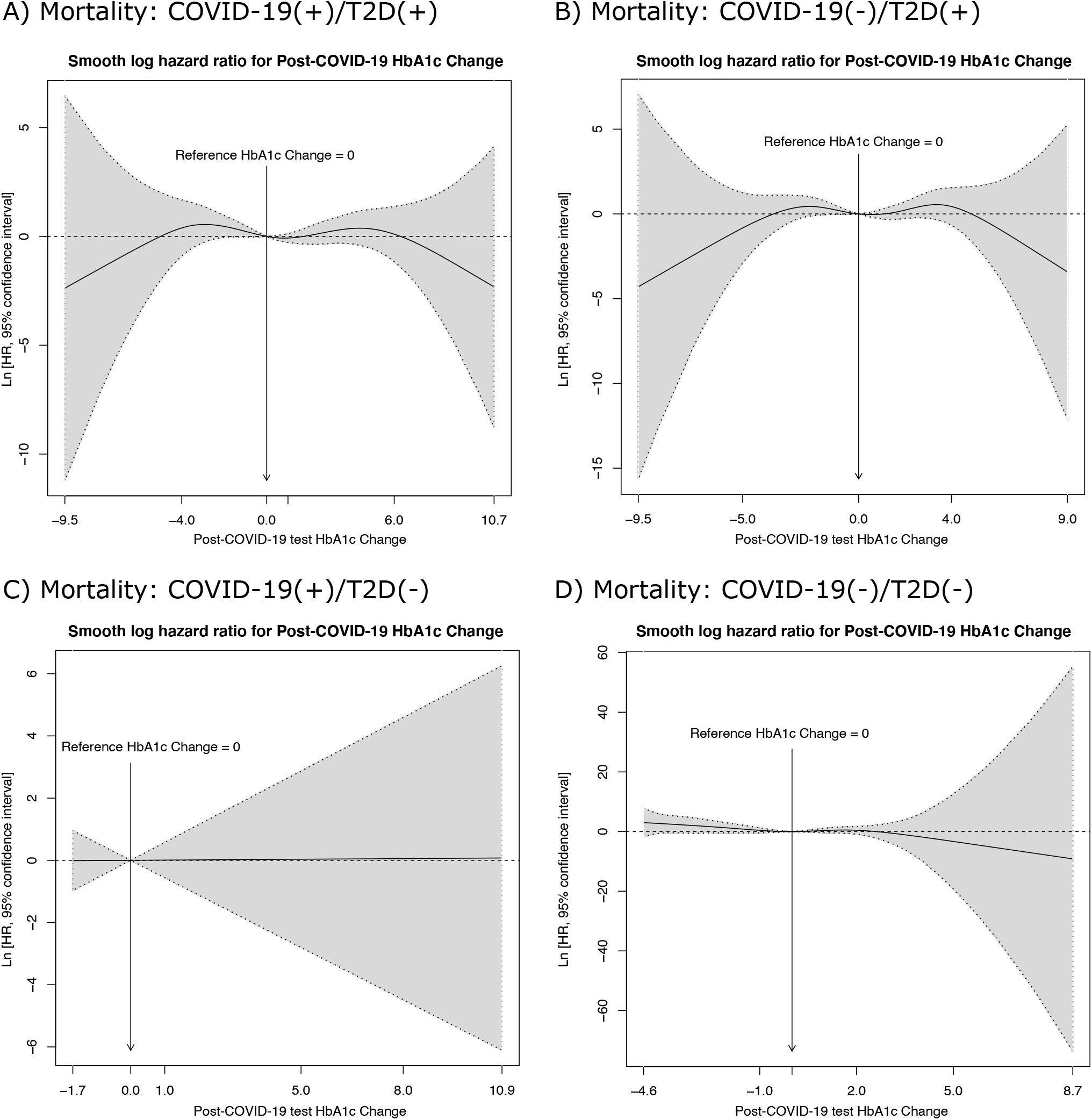
Non-parametric estimates of the dependence of risk of all-cause mortality on post-COVID-19 test HbA1c in (A) COVID-19(+) patients with T2D diagnosis, (B) COVID-19(-) patients with T2D diagnosis, (C) COVID-19(+) patients without T2D diagnosis, and (D) COVID-19(-) patients without T2D diagnosis. The reference HbA1c change was set to 0. The plots were generated using restricted cubic splines (rcs) implemented in Cox proportional hazard models using smoothHR package in R (2).

**Supplementary Figure 7:**
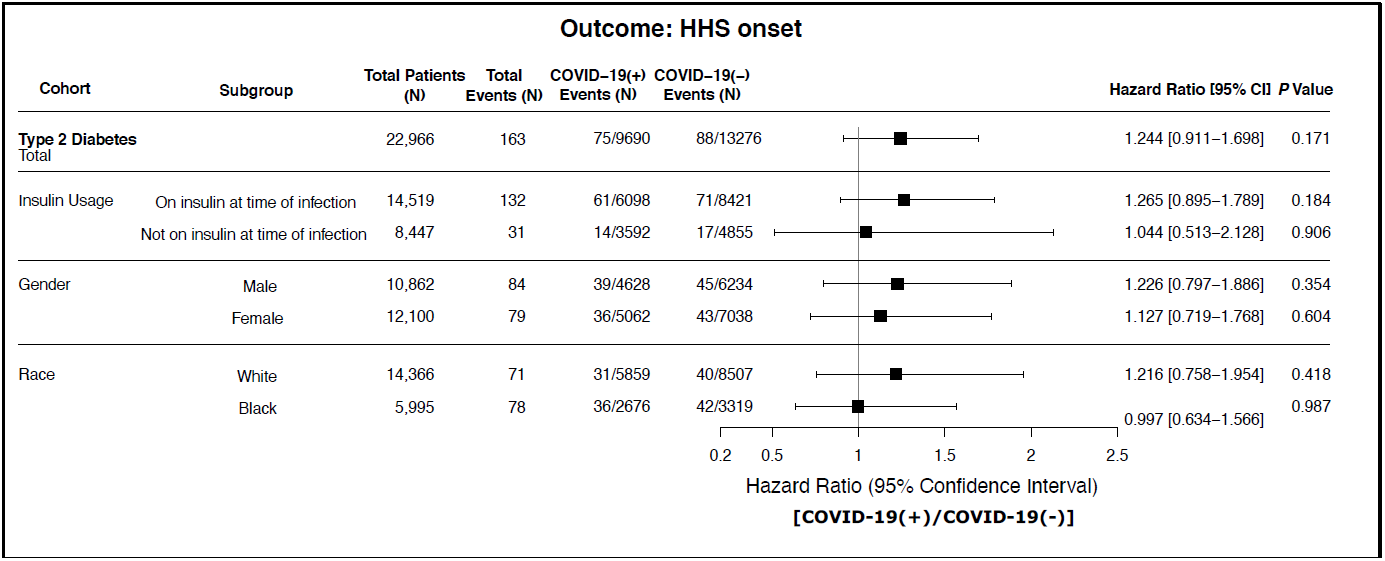
Association of risk of Hyperosmolar hyperglycemic syndrome (HHS) onset and COVID-19 test status in T2D(+) patients. The T2D(+) cohort was further split up based on baseline insulin usage. The patients with pre-existing HHS were removed from the T2D(+) cohort. The column COVID-19(+) events represent the proportion of number of [COVID-19(+)/HHS(+)]/[COVID-19(+)/HHS(+/-)] events. Similarly, the column COVID-19(-) events represent the proportion of number of [COVID-19(-)/HHS(+)]/[COVID-19(-)/HHS(+/-)] events.

